# Imaging-detected benign breast findings in a forensic autopsy cohort unselected for breast symptoms: descriptive results from the Sisyphus study

**DOI:** 10.64898/2026.05.07.26352434

**Authors:** Zacharoula Sidiropoulou, Carlos Santos

**Author notes:** **Corresponding Author:** Zacharoula Sidiropoulou, MD, PhD, Nova Medical School, Lisbon, Portugal.

## Abstract

**Rationale and Objectives:** Published estimates of benign breast disease (BBD) are derived mainly from clinical, surgical, screening-recall, or reduction-mammoplasty series. Forensic autopsy cohorts can reduce referral and symptom-selection bias, although they are not necessarily representative of the whole living population. We describe imaging-detected benign breast findings in the Sisyphus forensic autopsy cohort.

**Materials and Methods:** Consecutive medico-legal autopsies of individuals aged 40 years or older were prospectively evaluated over a multi-year period at a medico-legal autopsy service in Portugal. Bilateral breast specimens obtained by subcutaneous modified radical mastectomy were examined with specimen digital mammography and ultrasonography. Findings were classified according to BI-RADS terminology. Lesions requiring tissue diagnosis in the post-mortem protocol underwent wire-guided or direct excisional biopsy. Female cadavers were analysed as the primary cohort; male cadavers were analysed separately as an exploratory subgroup. Proportions are reported with exact 95% confidence intervals (CIs).

**Results:** The cohort included 291 cadavers: 217 women and 74 men. Among female breast specimens, 236/434 were BI-RADS 1 (54.4%; 95% CI, 49.6-59.1), 189/434 were BI-RADS 2 (43.5%; 95% CI, 38.8-48.4), and 8/434 were protocol-sampled suspicious findings (1.8%; 95% CI, 0.8-3.6). At the cadaver level, 99/217 women had at least one benign imaging finding (45.6%; 95% CI, 38.9-52.5). Mammographic benign findings were present in 91/217 women (41.9%; 95% CI, 35.3-48.8), dominated by calcifications; ultrasonographic benign findings were present in 51/217 (23.5%; 95% CI, 18.0-29.7), most often simple cysts and duct ectasia. Plasma cell mastitis-pattern calcifications were observed in 8/217 women (3.7%; 95% CI, 1.6-7.1). Male benign findings were less frequent (9/74, 12.2%; 95% CI, 5.7-21.8) and were dominated by benign lymph-node variants. All nine protocol-sampled lesions were benign at histology. Clinical breast examination identified 5/8 protocol-sampled female lesions (62.5%; 95% CI, 24.5-91.5).

**Conclusion:** In this forensic autopsy cohort unselected for breast symptoms, benign imaging findings were common in women aged 40 years or older and less frequent in men. The results provide descriptive post-mortem imaging reference data, but lesion-specific estimates, especially rare entities, should be interpreted with caution because of small numerators, the older age profile, limited clinical history, and the original cancer-focused design of the Sisyphus study.

## 1. Introduction

Benign breast disease (BBD) comprises a heterogeneous group of non-malignant breast findings, including cysts, duct ectasia, fibrocystic changes, fibroadenoma, fat necrosis, benign calcifications, inflammatory conditions, and other proliferative and non-proliferative lesions. These entities are common in breast imaging practice and frequently drive additional diagnostic assessment, short-interval follow-up, or biopsy.

Reliable imaging-based estimates of BBD are difficult to obtain. Most published data come from surgical biopsy registries, screening-recall cohorts, symptomatic breast clinics, or reduction-mammoplasty specimens. These sources answer clinically important questions, but they are also enriched for selected symptoms, mammographic abnormalities, tissue excision, breast size, or age groups. As a result, they do not fully describe the background imaging spectrum of benign findings in individuals who were not selected because of breast symptoms or screening recall.

Forensic autopsy cohorts offer a complementary approach. When breast tissue is retrieved systematically from individuals who died unexpectedly and were not selected for breast disease, imaging findings can be evaluated outside the usual clinical referral pathway. This design reduces several forms of clinical selection bias, but it does not make the cohort automatically representative of the general population. Age, cause of death, forensic inclusion criteria, regional demographics, and the post-mortem setting must all be considered when interpreting such data.

The Sisyphus study was originally designed to evaluate imaging-detected silent breast cancer in autopsy specimens. The primary cancer analyses have been published previously. The same study, however, generated a systematic mammographic and ultrasonographic dataset of benign breast findings in both sexes. The present manuscript reports those benign findings as a dedicated descriptive analysis. The objectives were to describe the spectrum of imaging-detected benign breast findings in women, report male findings separately as an exploratory subgroup, provide exact confidence intervals for prevalence estimates, and clarify the limitations inherent to post-mortem breast imaging.

## 2. Materials and Methods

### 2.1 Study design and population

The Sisyphus study was a prospective, consecutive medico-legal autopsy investigation performed over a multi-year period at a medico-legal autopsy service in Portugal, in collaboration with a hospital breast-imaging unit. Ethics approval was granted by the Ethics Committee for Health of Centro Hospitalar de Lisboa Ocidental (approval number 20170700050), and the study was registered at ClinicalTrials.gov (NCT02480933).

Cadavers aged 40 years or older who met medico-legal inclusion criteria and for whom breast specimen retrieval was authorised were eligible. Cases were excluded when the post-mortem interval exceeded 48 hours, when extensive trauma or decomposition affected one or both breasts, or when known or clinically evident breast cancer was present. The current analysis focuses on benign imaging findings; malignant cases identified or excluded in the primary cancer-focused Sisyphus analyses were not included in the benign prevalence tables. This design choice isolates benign findings but may underestimate the frequency of benign proliferative lesions that coexist with malignancy.

Female and male cadavers were analysed separately because the anatomy, disease spectrum, and clinical interpretation of male and female breast imaging differ substantially. Female cadavers constituted the primary analytic cohort. Male cadavers were retained as an exploratory subgroup because systematic imaging-based post-mortem data on benign male breast findings are scarce.

### 2.2 Specimen collection and imaging protocol

After eligibility confirmation, bilateral subcutaneous modified radical mastectomy (bsMRM) was performed through a Dufourmentel incision in fresh cadavers. Specimens were oriented, sealed, transported to the collaborating hospital breast-imaging unit, and evaluated within the early post-mortem interval. The mean interval from death to biopsy was approximately 18 hours.

Each specimen underwent measurement, weighing, clinical breast examination by inspection and palpation by a dedicated breast surgeon, specimen digital mammography, and breast ultrasonography. Ultrasonography was performed using a GE Healthcare LOGIQ S7 Expert system with a 9-15 MHz transducer. Digital mammography was performed using a GE Healthcare Senographe Essential system at 27 kV, 60-70 mA, and 10-15 daN compression.

All cadavers included in the imaging analysis underwent specimen mammography and ultrasonography. Therefore, statements such as ‘mammographic findings were recorded in 91 of 217 female cadavers’ refer to positive mammographic findings; the remaining imaged cadavers had no recorded mammographic benign abnormality.

### 2.3 Imaging classification and post-mortem tissue sampling

Findings were described using the American College of Radiology BI-RADS 5th edition lexicon. Benign calcifications were recorded when their morphology and distribution fulfilled BI-RADS benign criteria, including typical coarse, vascular, scattered, or lucent-centered morphology. Isolated non-reproducible punctate calcifications without a defined benign or suspicious pattern were not counted as a positive finding.

The post-mortem design required separation of imaging interpretation from practical tissue-sampling management. In living patients, BI-RADS 3 findings are usually managed by short-interval follow-up rather than immediate biopsy. Because interval follow-up is impossible in cadaver studies, lesions considered to require confirmation under the study protocol were sampled immediately. This practical decision should not be interpreted as changing the intrinsic BI-RADS meaning of a probably benign lesion or as creating a clinical positive predictive value for BI-RADS 4a. For that reason, the present revision reports sampled lesions descriptively and does not use malignancy positive predictive value as a primary endpoint.

Wire-guided biopsy was used when the target required radiologic localisation within the specimen. Direct excisional biopsy was used when the lesion was palpable, externally identifiable, or sufficiently localised by specimen orientation and imaging coordinates to permit targeted excision without wire placement. All sampled tissue was submitted for histopathology.

### 2.4 Histopathology

Biopsy specimens were fixed in 10% buffered formalin, paraffin-embedded, processed routinely, and sectioned at 3 micrometres. Immunocytochemistry for oestrogen receptor, progesterone receptor, and Ki-67 was performed when indicated. Slides were reviewed by an experienced breast pathologist.

### 2.5 Statistical analysis

Proportions were calculated at the cadaver level for person-based findings and at the specimen level for BI-RADS classification. Exact 95% CIs were calculated using the Clopper-Pearson method. Age distribution was analysed by decade. Chi-square testing was used to compare broad BI-RADS distributions by sex and to explore age-decade variation in female benign findings.

Associations between BMI and BI-RADS classification were considered exploratory. The source dataset did not include detailed reproductive history, menopausal status, medication exposure, hormone therapy, or complete ante-mortem symptom history. Therefore, BMI correlations are reported descriptively and should not be interpreted as independent causal associations. Multivariable modelling of lesion-specific outcomes was not attempted because several entities had small numerators.

### 2.6 Contextual literature comparison

Published benchmarks were used only to contextualise the observed range of findings and were not treated as formal meta-analytic comparators. Differences in age structure, imaging technique, symptomatic status, screening practices, and histologic confirmation limit direct comparison between the Sisyphus cohort and clinical series.

## 3. Results

### 3.1 Cohort characteristics

The cohort included 291 cadavers: 217 women and 74 men. The female cohort had a mean age of 65.5 years, and the male cohort had a mean age of 63.9 years. All cadavers were aged 40 years or older. Mean BMI was 24.9 kg/m2 in women and 28.6 kg/m2 in men. Acute myocardial infarction was the most common cause of death in both sexes in the previously reported Sisyphus cohort. Cause-of-death categories were not used as predictors in the current benign imaging analysis.

No included cadaver had a known or clinically evident breast cancer diagnosis at the time of autopsy. Detailed ante-mortem breast symptoms, family history, hormone exposure, reproductive history, prior benign breast diagnoses, and menopausal status were not available for systematic analysis.

### 3.2 Female cohort: overall imaging classification

Among 434 female breast specimens, 236 were BI-RADS 1 (54.4%; 95% CI, 49.6-59.1), 189 were BI-RADS 2 (43.5%; 95% CI, 38.8-48.4), and 8 underwent protocol tissue sampling for suspicious or follow-up-ineligible findings (1.8%; 95% CI, 0.8-3.6). At the cadaver level, 99 of 217 women had at least one benign imaging finding (45.6%; 95% CI, 38.9-52.5). The broad distribution of negative, benign, and protocol-sampled findings varied significantly by sex (chi-square p < 0.001), reflecting the lower frequency and different spectrum of male findings.

### 3.3 Female mammographic findings

All 217 female cadavers underwent specimen mammography. Positive mammographic benign findings were recorded in 91 women (41.9%; 95% CI, 35.3-48.8). Microcalcifications of any recorded type were identified in 42 women (19.4%; 95% CI, 14.3-25.2), including dispersed microcalcifications in 35 women (16.1%; 95% CI, 11.5-21.7) and localised microcalcifications in 7 women (3.2%; 95% CI, 1.3-6.5).

Macrocalcifications alone were present in 13 women (6.0%; 95% CI, 3.2-10.0), while concurrent microcalcifications and macrocalcifications were present in 28 women (12.9%; 95% CI, 8.7-18.1).

A plasma cell mastitis-pattern of rod-like or periductal calcifications was observed incidentally in 8 women (3.7%; 95% CI, 1.6-7.1). Because of the small numerator, this finding is best interpreted as an observation in an autopsy imaging cohort rather than as a precise population prevalence estimate.

### 3.4 Female ultrasonographic findings

Ultrasonographic benign findings were recorded in 51 of 217 female cadavers (23.5%; 95% CI, 18.0-29.7). Simple cysts were recorded in 14 women (6.5%; 95% CI, 3.6-10.6), duct ectasia in 13 (6.0%; 95% CI, 3.2-10.0), and concurrent cysts and duct ectasia in 8 (3.7%; 95% CI, 1.6-7.1).

Less frequent observations included benign axillary lymph nodes in 6 women (2.8%; 95% CI, 1.0-5.9), fat necrosis in 5 (2.3%; 95% CI, 0.8-5.3), lipomas in 3 (1.4%; 95% CI, 0.3-4.0), and intramammary lymph nodes in 2 (0.9%; 95% CI, 0.1-3.3). These rare lesion-specific estimates should be interpreted cautiously because their confidence intervals are wide.

### 3.5 Age-decade distribution in women

After correcting the allocation of protocol-sampled cases to age decades, the proportion of women with any benign imaging finding ranged from 37.8% in the oldest group to 51.1% in the 70-79-year group. The age-decade trend did not reach statistical significance (chi-square = 1.77, df = 4, p = 0.779). Because the cohort was restricted to individuals aged 40 years or older and was predominantly post-menopausal, these data should not be generalised to younger reproductive-age women.

### 3.6 BMI and imaging category

Exploratory Spearman analysis showed a weak positive correlation between BMI and higher imaging category in women (r_s = 0.143, p = 0.031) and a weak inverse correlation in men (r_s = -0.182, p = 0.028). Weight was also inversely associated with imaging category in men (r_s = -0.202, p = 0.020). Because age, menopausal status, breast density, hormone exposure, symptoms, and other possible confounders were not available for complete multivariable adjustment, these associations are reported as hypothesis-generating only.

### 3.7 Male exploratory findings

Among 148 male breast specimens, 129 were BI-RADS 1 (87.2%; 95% CI, 80.7-92.1), 18 were BI-RADS 2 (12.2%; 95% CI, 7.4-18.5), and 1 underwent protocol tissue sampling (0.7%; 95% CI, 0.0-3.7). At the cadaver level, 9 of 74 men had at least one benign imaging finding (12.2%; 95% CI, 5.7-21.8).

The male imaging spectrum differed from the female spectrum. Findings were dominated by benign lymph-node variants and occasional calcifications rather than by cysts, duct ectasia, or other typically female benign entities. The single male protocol-sampled lesion showed reactive intramammary lymph nodes without neoplasia. Gynecomastia was not a prespecified endpoint of the original cancer-focused dataset; therefore, this study cannot estimate gynecomastia prevalence reliably.

### 3.8 Protocol-sampled lesions and histology

Nine lesions underwent tissue sampling: eight in women and one in a man. All were benign at histology. Female histologic diagnoses included fibrocystic changes, fat necrosis, simple fibroadenoma, calcified fibroadenoma with intraductal microcalcification, partially calcified microcyst, and hamartoma. The male biopsy showed reactive lymph nodes without neoplasia. Exact ages, case identifiers, lesion sizes, and individual-level combinations of age, sex, imaging target, and histology were removed from the revised table to reduce re-identification risk. Because known or detected malignant cases were excluded from this benign analysis and because post-mortem sampling was influenced by the impossibility of interval follow-up, malignancy positive predictive value was not calculated.

### 3.9 Clinical breast examination

Among the eight protocol-sampled female lesions, clinical breast examination identified five and missed three, corresponding to an apparent sensitivity of 62.5% (95% CI, 24.5-91.5) and an apparent false-negative proportion of 37.5% (95% CI, 8.5-75.5) in this limited setting. One abnormal clinical examination had no corresponding imaging finding. These estimates are based on very small numbers and on post-mortem specimen examination; they should not be directly extrapolated to living clinical practice.

## 4. Discussion

This revised analysis describes benign breast imaging findings in a forensic autopsy cohort unselected for breast symptoms. The principal observation is that benign imaging findings were common among women aged 40 years or older, while male findings were substantially less frequent and qualitatively different. Exact CIs show that common categories such as any benign finding or any mammographic finding are estimated with moderate precision, whereas rare entities such as plasma cell mastitis-pattern calcifications, fat necrosis, lipomas, and intramammary lymph nodes have wide CIs and should be interpreted as observations rather than definitive population prevalence estimates.

The female findings were dominated by mammographic calcifications and by common ultrasonographic benign entities such as simple cysts and duct ectasia. This pattern is biologically plausible in a cohort with a mean age in the mid-60s, where involutional change, calcified benign lesions, vascular or stromal calcification, and post-menopausal regression of cystic disease are expected. The observed cyst frequency was at the lower end of published clinical ranges, which may reflect the older age structure and the exclusion of younger reproductive-age women.

Plasma cell mastitis-pattern calcifications were detected incidentally in a small number of women. Although the numerator was low, the finding is clinically relevant because periductal rod-like or lucent-centered calcifications can create diagnostic uncertainty in screening practice. The post-mortem setting indicates that such patterns may exist without an ante-mortem clinical diagnosis. However, the estimate should not be presented as a precise prevalence rate without a larger cohort.

The male subgroup is potentially useful but must remain exploratory. Male benign breast imaging is clinically different from female breast imaging, and the present cohort was not designed specifically to characterise male breast disease. The predominance of lymph-node variants in men may partly reflect lower tissue volume and greater conspicuity of intramammary or axillary nodes, particularly in leaner individuals. Nevertheless, the absence of a prespecified gynecomastia endpoint and the small number of positive male cases prevent broader conclusions.

The clinical breast examination findings should also be interpreted cautiously. The study setting removed some barriers present in living clinical practice, such as patient discomfort or time pressure, but introduced others, including post-mortem tissue change, specimen handling, absence of patient-reported symptoms, and lack of dynamic clinical interaction. The apparent false-negative proportion among sampled lesions supports the complementary value of imaging, but it does not provide a definitive estimate of CBE performance in living screening populations.

Several methodological issues are specific to post-mortem imaging. Early autolysis, rigor mortis, tissue dehydration, post-mortem fluid redistribution, specimen compression, and bsMRM-related distortion could influence echogenicity, mammographic density, lesion conspicuity, and palpation. The early post-mortem imaging interval and systematic protocol reduce but do not eliminate these concerns. Future work should include formal radiologic-pathologic validation of post-mortem imaging appearance across defined time intervals.

The study has important strengths. It was prospective and consecutive, used systematic bilateral mammography and ultrasonography, applied a breast-imaging pathway to specimens from individuals not selected for breast symptoms, and included both women and men. It also provides exact CIs and separates female and male analyses. These features make the dataset a useful descriptive reference for post-mortem breast imaging.

Limitations are equally important. The cohort was single-centre, regionally specific, and older, with no participants younger than 40 years. It was originally powered and designed for cancer detection, not for benign lesion-specific prevalence. Detailed ante-mortem history, symptoms, hormonal exposure, reproductive factors, menopausal status, family history, and prior benign breast diagnoses were unavailable. BMI data were incomplete in the source study and could not be fully adjusted for confounding. Histologic confirmation was performed only for protocol-sampled lesions, so most BI-RADS 2 findings rely on established imaging criteria rather than tissue diagnosis. Finally, exclusion of malignant cases may reduce the observed burden of benign proliferative lesions associated with cancer.

## 5. Conclusion

In this prospective forensic autopsy cohort unselected for breast symptoms, benign imaging findings were common in women aged 40 years or older and substantially less frequent in men. Female findings were dominated by benign calcifications, cysts, duct ectasia, and other involutional or non-malignant entities, while male findings were mainly benign lymph-node variants. The study provides descriptive reference data for systematic post-mortem mammography and ultrasonography, but its estimates should be interpreted in light of wide CIs for rare findings, an older age distribution, absence of detailed ante-mortem history, possible post-mortem imaging effects, and the original cancer-focused design of the Sisyphus study.

**Table 1.** Cohort characteristics.

| Sex | n cadavers | Mean age (years) | Eligibility age range | Mean BMI (kg/m2) | Analytic role |
| --- | --- | --- | --- | --- | --- |
| Women | 217 | 65.5 | >=40 years | 24.9 | Primary analytic cohort |
| Men | 74 | 63.9 | >=40 years | 28.6 | Exploratory subgroup |
| Total | 291 | - | >=40 years | - | Consecutive medico-legal autopsy cohort |

BMI, body mass index. Exact individual ages are not reported. Detailed ante-mortem breast symptoms, hormone exposure, reproductive history, and menopausal status were not available for systematic analysis.

**Table 2.**
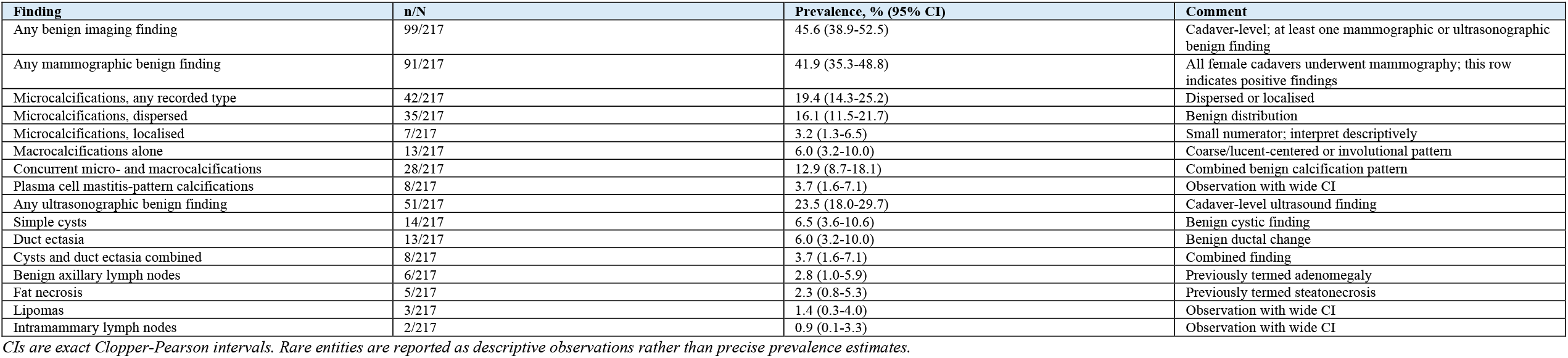
Female cadaver-level benign imaging findings.

**Table 3.** Specimen-level imaging classification by sex.

| Sex | Imaging category | n/N specimens | Percentage, % (95% CI) |
| --- | --- | --- | --- |
| Women | BI-RADS 1 negative | 236/434 | 54.4 (49.6-59.1) |
| Women | BI-RADS 2 benign | 189/434 | 43.5 (38.8-48.4) |
| Women | Protocol-sampled/suspicious | 8/434 | 1.8 (0.8-3.6) |
| Men | BI-RADS 1 negative | 129/148 | 87.2 (80.7-92.1) |
| Men | BI-RADS 2 benign | 18/148 | 12.2 (7.4-18.5) |
| Men | Protocol-sampled/suspicious | 1/148 | 0.7 (0.0-3.7) |

The category 'protocol-sampled/suspicious' reflects lesions sampled in the post-mortem protocol. It should not be interpreted as a clinical BI-RADS 4 positive predictive value.

**Table 4.** Female benign imaging findings by age decade.

| Age decade | n women | BI-RADS 2 / benign imaging n | BI-RADS 2 / benign imaging % (95% CI) | Protocol-sampled n | Any benign n | Any benign % (95% CI) |
| --- | --- | --- | --- | --- | --- | --- |
| 40-49 | 33 | 12 | 36.4 (20.4-54.9) | 2 | 14 | 42.4 (25.5-60.8) |
| 50-59 | 41 | 18 | 43.9 (28.5-60.3) | 2 | 20 | 48.8 (32.9-64.9) |
| 60-69 | 59 | 27 | 45.8 (32.7-59.2) | 0 | 27 | 45.8 (32.7-59.2) |
| 70-79 | 47 | 21 | 44.7 (30.2-59.9) | 3 | 24 | 51.1 (36.1-65.9) |
| >=80 | 37 | 13 | 35.1 (20.2-52.5) | 1 | 14 | 37.8 (22.5-55.2) |
| Total | 217 | 91 | 41.9 (35.3-48.8) | 8 | 99 | 45.6 (38.9-52.5) |

Rows are cadaver-level summaries. Protocol-sampled counts were corrected to align with the de-identified source allocation. Age-decade trend for any benign finding: chi-square = 1.77, df = 4, p = 0.779.

**Table 5.**
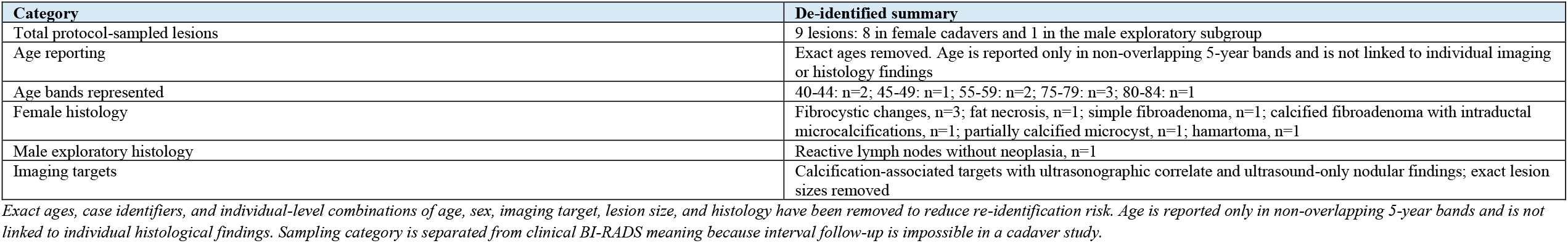
Summary of all protocol-sampled lesions.

| Category | De-identified summary |
| --- | --- |
| Total protocol-sampled lesions | 9 lesions: 8 in female cadavers and 1 in the male exploratory subgroup |
| Age reporting | Exact ages removed. Age is reported only in non-overlapping 5-year bands and is not linked to individual imaging or histology findings |
| Age bands represented | 40-44: n=2; 45-49: n=1; 55-59: n=2; 75-79: n=3; 80-84: n=1 |
| Female histology | Fibrocystic changes, n=3; fat necrosis, n=1; simple fibroadenoma, n=1; calcified fibroadenoma with intraductal microcalcifications, n=1; partially calcified microcyst, n=1; hamartoma, n=1 |
| Male exploratory histology | Reactive lymph nodes without neoplasia, n=1 |
| Imaging targets | Calcification-associated targets with ultrasonographic correlate and ultrasound-only nodular findings; exact lesion sizes removed |

Exact ages, case identifiers, and individual-level combinations of age, sex, imaging target, lesion size, and histology have been removed to reduce re-identification risk. Age is reported only in non-overlapping 5-year bands and is not linked to individual histological findings. Sampling category is separated from clinical BI-RADS meaning because interval follow-up is impossible in a cadaver study.

**Table 6.**
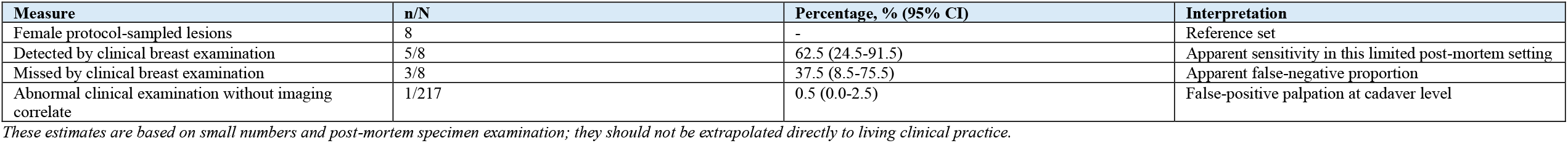
Clinical breast examination findings in the female cohort.

**Figure 1.**
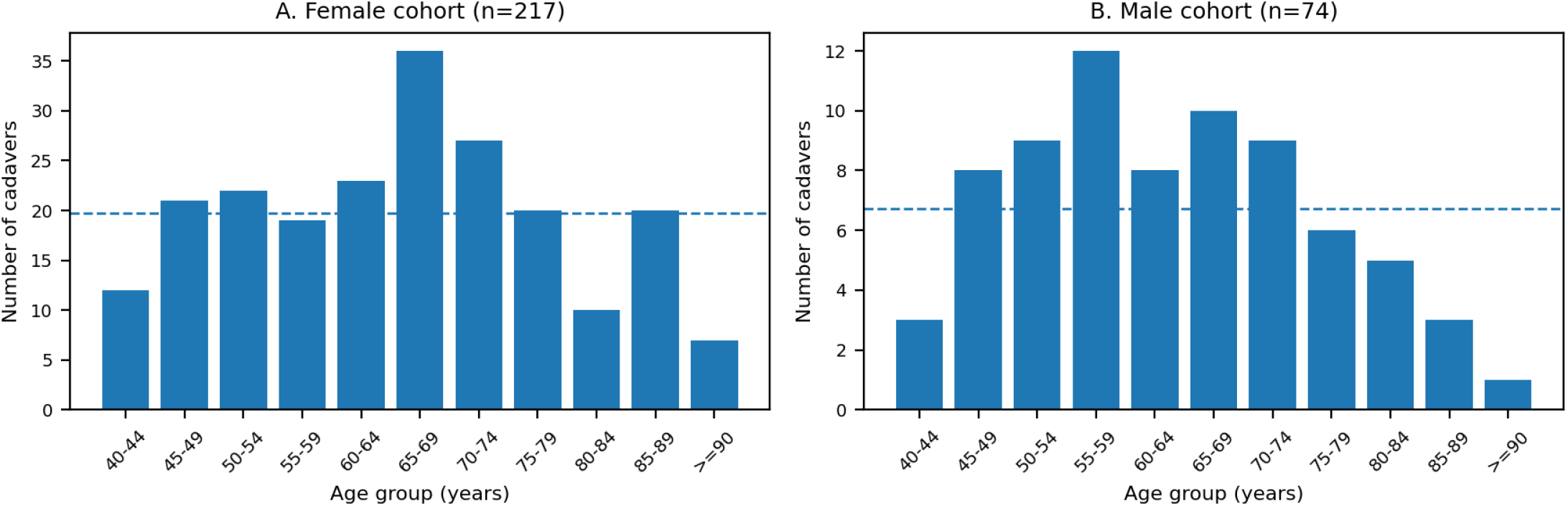
Age distribution of the Sisyphus forensic cohort by sex. Dashed lines indicate the mean number of cadavers per age bin.

**Figure 2.**
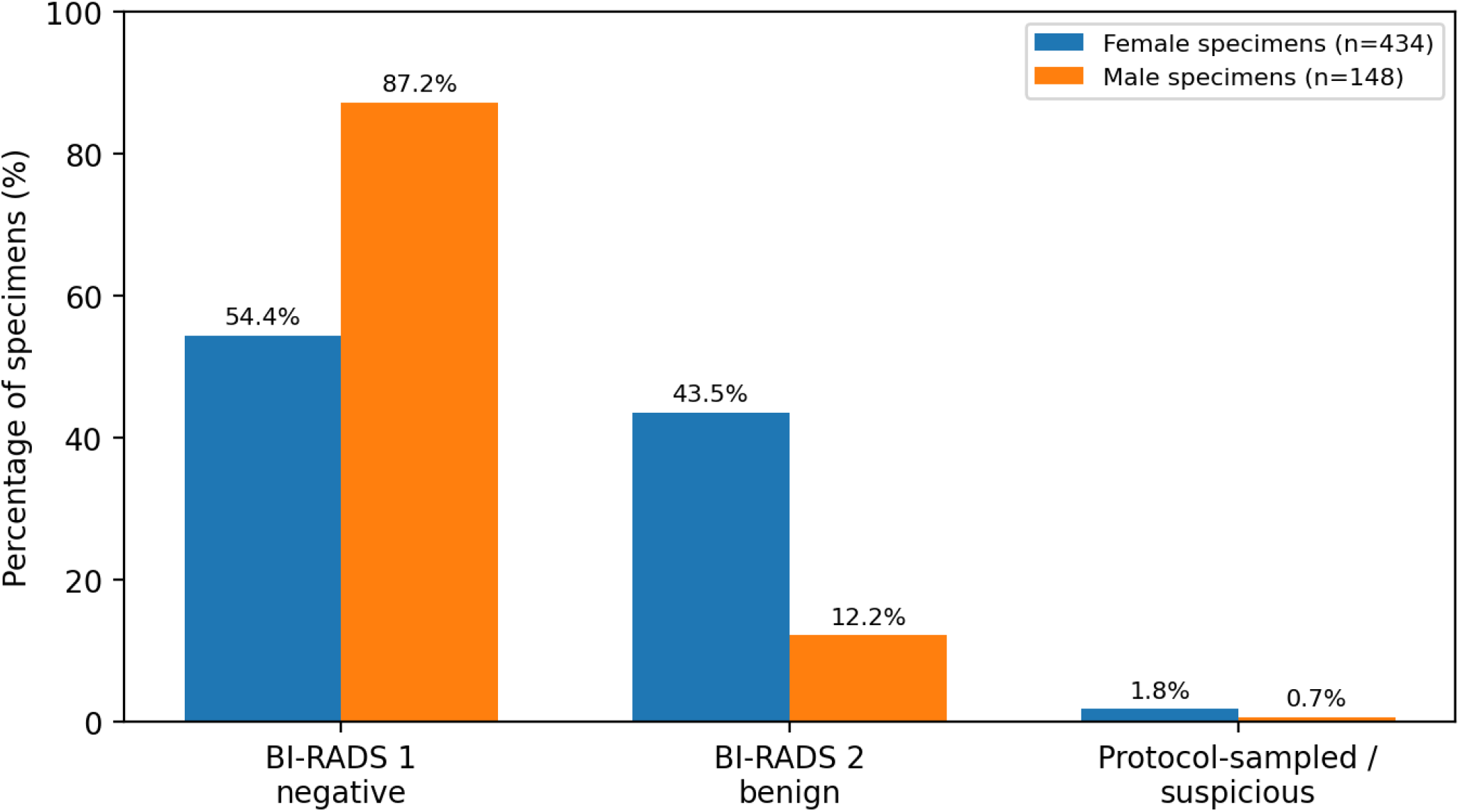
Specimen-level imaging classification by sex. The broad distribution differed between women and men (chi-square p < 0.001). The protocol-sampled/suspicious category reflects post-mortem tissue-sampling decisions and should not be interpreted as a clinical positive predictive value category.

**Figure 3.**
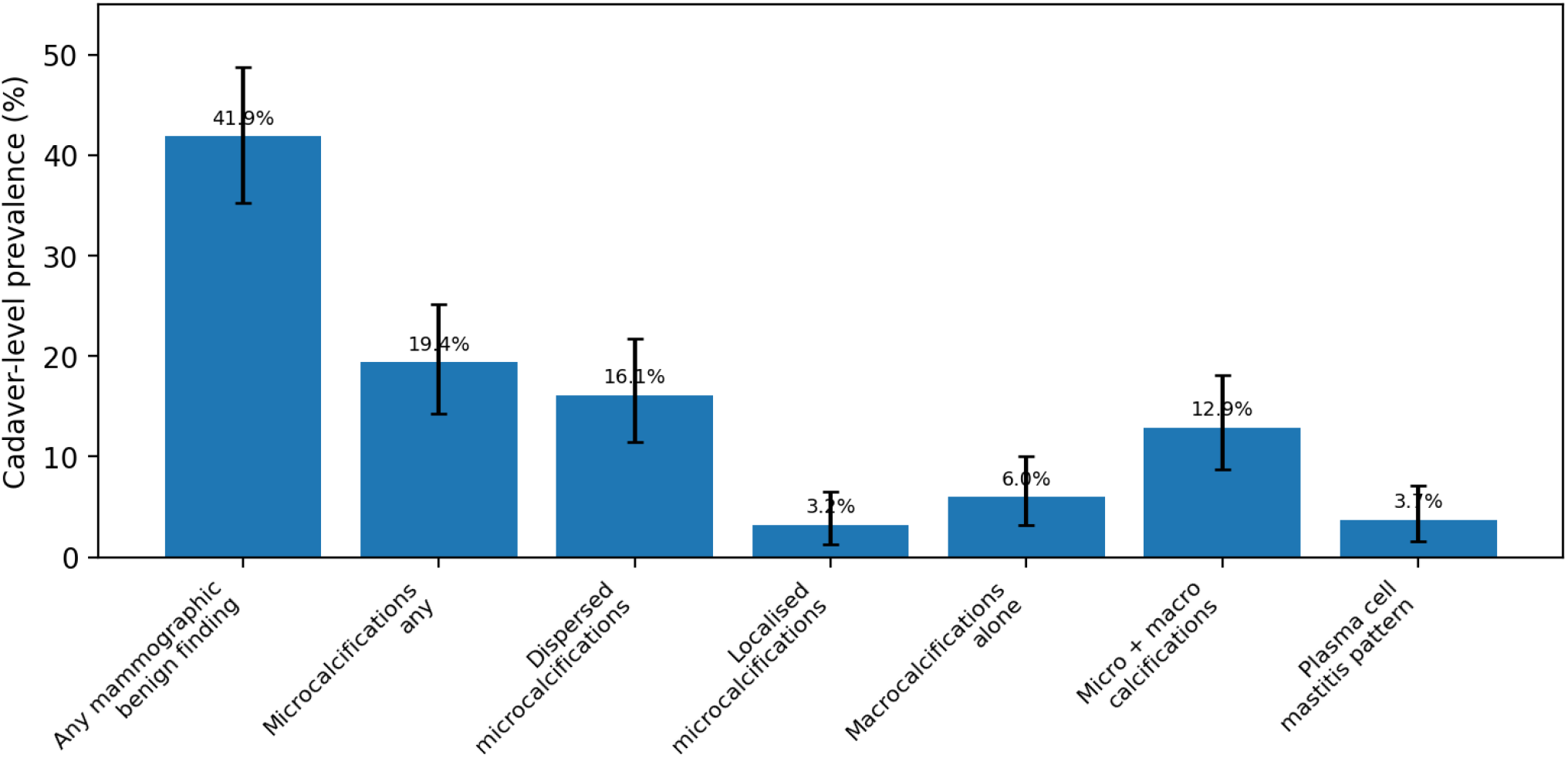
Mammographic benign findings in female cadavers. Bars show cadaver-level prevalence; error bars show exact 95% confidence intervals.

**Figure 4.**
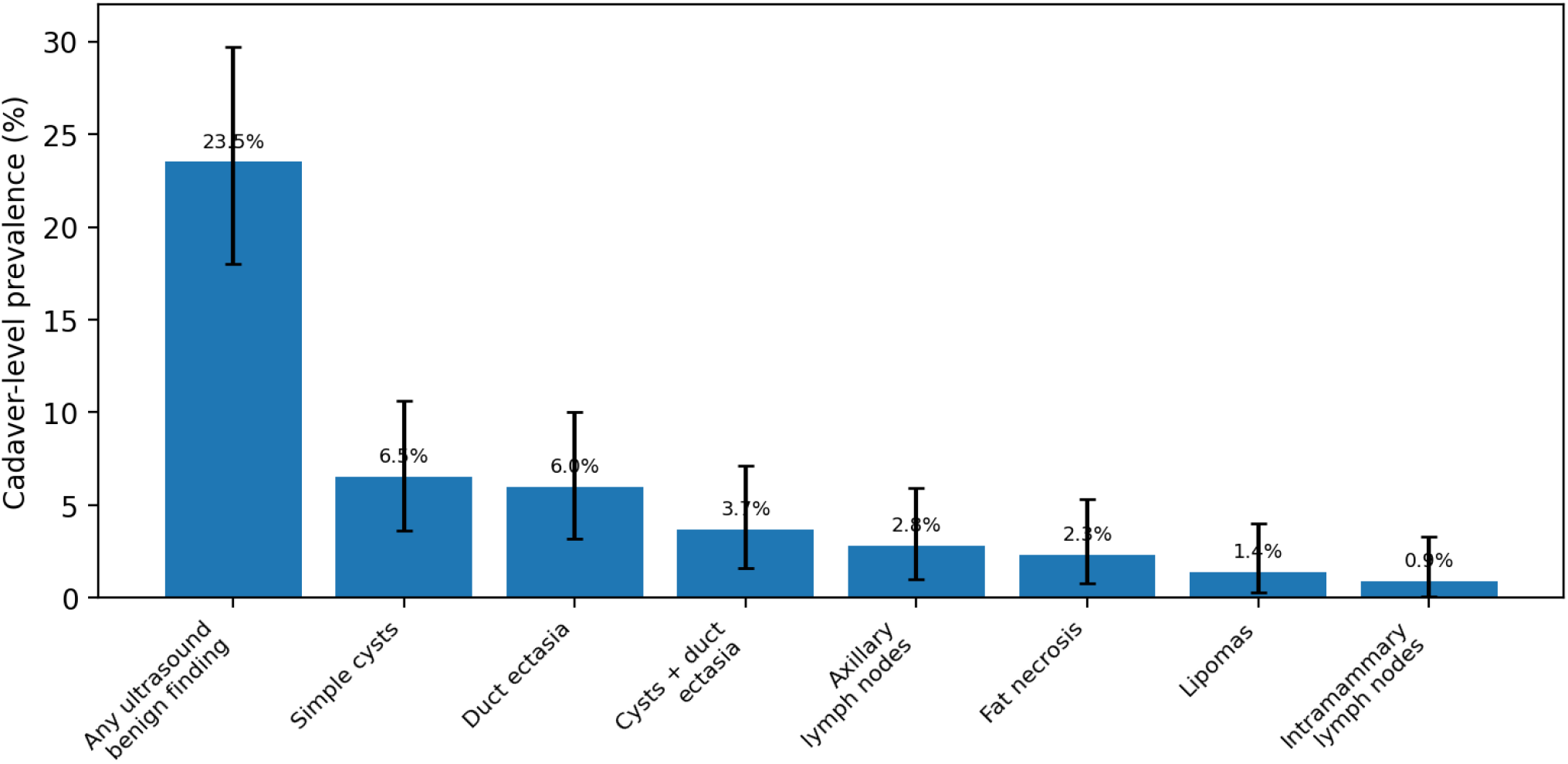
Ultrasonographic benign findings in female cadavers. Bars show cadaver-level prevalence; error bars show exact 95% confidence intervals.

**Figure 5.**
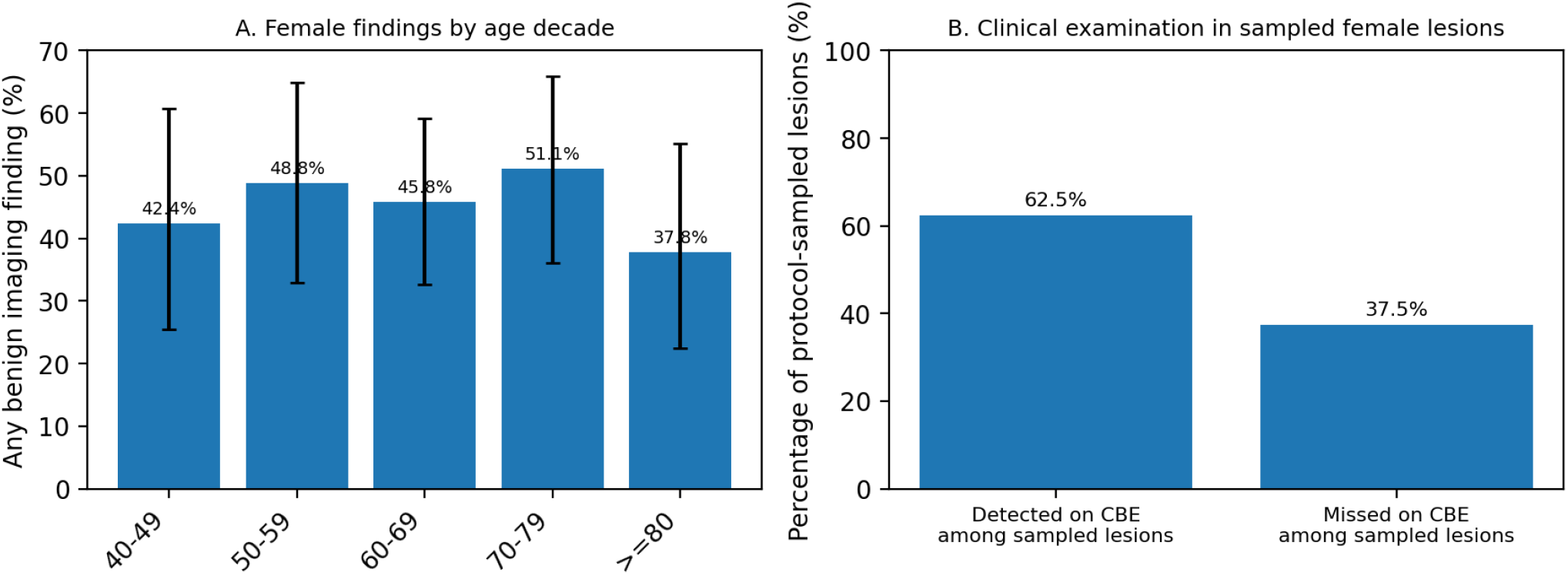
Female benign findings by age decade and clinical examination context. Error bars in panel A show exact 95% confidence intervals based on corrected age-decade allocation. Panel B is limited to the eight protocol-sampled female lesions.

## Data Availability

All data produced in the present study are available upon reasonable request to the authors

## Declarations

Ethics approval and registration: Ethics approval was granted by the Ethics Committee for Health of Centro Hospitalar de Lisboa Ocidental, approval number 20170700050. The study protocol was registered at ClinicalTrials.gov (NCT02480933).

### Data availability

Aggregate data supporting the findings are available from the corresponding author on reasonable request, subject to medico-legal restrictions.

Prior publications from this cohort: The primary cancer outcomes of the Sisyphus cohort have been reported previously. The present analysis reports benign imaging findings as a dedicated outcome.

## Author contributions

ZS: Conceptualisation, methodology, data curation, formal analysis, writing - original draft, writing - review and editing, project administration. CS: inclusion and exclusion criteria and autopsy authorisations.

## Abbreviations

BBD: benign breast disease
BI-RADS: Breast Imaging Reporting and Data System
BMI: body mass index
bsMRM: bilateral subcutaneous modified radical mastectomy
CBE: clinical breast examination
CI: confidence interval
DCIS: ductal carcinoma in situ
INMLCF: Instituto Nacional de Medicina Legal e Ciencias Forenses.

